# Antibody profiling of COVID-19 patients in an urban low-incidence region in Northern Germany

**DOI:** 10.1101/2020.05.30.20111393

**Authors:** Werner Solbach, Julia Schiffner, Insa Backhaus, David Burger, Ralf Staiger, Bettina Tiemer, Andreas Bobrowski, Timothy Hutchings, Alexander Mischnik

## Abstract

This explorative monocentric study shows IgA and IgG antibody profiles from 110 patients with self-reported mild to moderate, or no COVID-19 related symptoms after laboratory-confirmed infection with SARS-CoV-2. The study region is in an urban and well-defined environment in a low-incidence region in Northern Germany. We found that approx. 70 % of the patients developed sustainable antibodies 3 weeks or later after the infection. In about 30 % of the patients with mild to moderate symptoms, no significant antibodies could be detected in two consecutive analyses. Conversely, out of ten patients without symptoms, four were repeatedly positive. Expectedly, six had no specific antibodies. The data indicate that antibody-positivity is a useful indicator of a previous SARS-CoV-2 infection. Negative antibodies do not rule out SARS-CoV-2 infection. Future studies need to determine the functionality of the antibodies in terms of personal protection and ability to transmit the virus.

## Introduction

The novel severe acute respiratory syndrome coronavirus (SARS-CoV-2) causes a respiratory disease, known as COVID-19^1^. On December 31, 2019, Chinese officials reported a cluster of cases of pneumonia in Wuhan, China. The infection was quickly qualified as epidemic^2^. As of January 30, 2020 it was announced a public health emergency of international concern. As of March 11, 2020 WHO officially declared the epidemic a pandemic^3^. Early reports from China and Italy indicated that SARS-CoV-2 causes illness of varying degrees^4^. The infection can spread easily as the virus is able to transmit during the asymptomatic or presymptomatic phase of infection^5, 6^. A vast majority of COVID-19 cases presents as mild or moderate symptomatic infection ranging from sore throat, cough and fever to radiographic pneumonia, particularly reported in children^7, 8^.

The first case in Germany was notified on January 27, 2020 and spread rapidly around the country^9, 10^. Germany’s national Public Health Institute (Robert Koch Institute [RKI]) on February 28, 2020 rated the risk of the COVID-19 pandemic for the population in Germany as “low to moderate”, since March 17 as “high” and for risk groups as “very high” since March 26^11^.

The core basis for the management of the outbreak is the early detection of SARS-CoV-2 in respiratory specimens (nasopharyngeal swabs) from patients presenting with clinical signs such as fever, dry cough or shortness of breath or in asymptomatic persons with close contacts to patients. People who have a cumulative face-to-face contact with a confirmed case for ≥15 min, direct contact with secretions or body fluids of a patient with confirmed COVID-19 disease, or, in the case of healthcare workers, work within 2 m of a patient with confirmed COVID-19 disease without personal protective equipment are at high risk for infection^12^.

The gold standard for SARS-CoV-2-detection is a specific polymerase reaction testing from a nasopharyngeal swab, sputum or broncoalveolar lavage^13^. Recently, commercial assays for serological analysis of specific COVID-19 antibodies became available^14, 15^. From a public health perspective an easy to establish and cost effective laboratory-based screening strategy may assist in rapid case detection, surveillance and ultimately in a better understanding of this epidemic^12^. Since there is no specific medical treatment or a vaccine available at present, it is crucial that sufficient herd immunity will develop in the population in order to interrupt uncontrollable transmission of the virus. Like in other coronaviruses, it is likely that neutralizing antibodies are central to the development of herd immunity to SARS-CoV-2. Therefore, insight into the development of immunity is pertinent for future guidance of preventive measures. In addition, antibody levels may give information on the question, whether patients with COVID-19 infection are immune to re-infection and/or can transmit the virus after recovery.

At present, different investigations are ongoing to get insight in seroprevalence of COVID-19 infection in Germany and Europe. Many researchers report from hot spot areas in Europe^16^ or regions of high prevalence in Germany^17^. The extent, duration and the protective function of the antibody response are not clear. Reports indicate that even among healthcare workers of a tertiary hospital with contact to COVID-19 patients seroprevalence is below 2 %^18^. Fortunately, the infection rate in northern Germany has been milder than in other parts of the country. Thanks to rigorous containment measures and early contact tracing, the Luebeck region always had incidence numbers less than the average of the rest of Germany.

In the present study, we aimed to explore the dynamics of the antibody response with respect to onset, level and duration in patients with confirmed SARS-CoV-2 infection in this low incidence region. Most of the study patients were outpatients with either mild, moderate or even without symptoms. The precise knowledge of the disease severity allowed us to attempt the clinical validation of the antibody development.

We determined serum antibody levels (immunoglobulin A and G) from 110 out of a total of 162 patients (as of April25) that qualified as COVID-19 index patients on the basis of clinical symptoms and concordant laboratory results for SARS-CoV-2 by polymerase chain reaction (PCR) according to the RKI case definition^19^. When first-degree contacts of index patients with or without symptoms proved to be SARS-CoV-2 positive by PCR, they were categorized as index patients and were included in the study. The disease severity of all patients was manageable by doctors in general practice or as outpatients in the local clinics. According to official rules, patients were quarantined for at least 14 days and two subsequent days without clinical symptoms. Sera were analyzed at various times after the end of the 14-days quarantine period.

## Materials and methods

### Study population and participant recruitment

The city of Luebeck (population 220,238) is situated in Northern Germany. The first two laboratory-confirmed SARS-CoV-2 cases were notified on February 29, 2020. The epidemic grew to 162 cases from which 151 recovered and one person (79 years) died (as of April 26). The incidence was 70.38 cases/100.000 inhabitants as compared to 178.33 for Germany (Figure 1).

**Figure 1.**
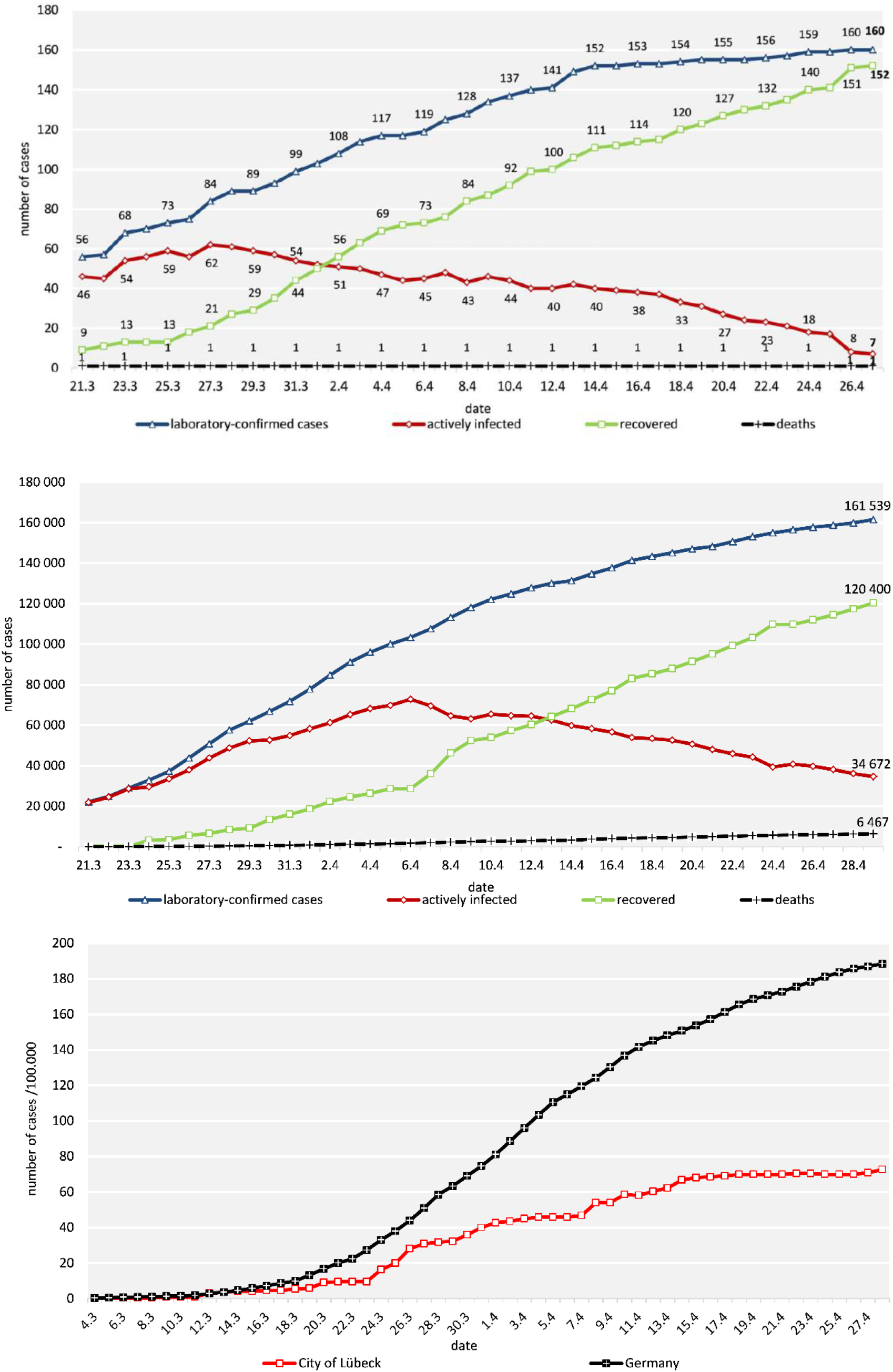
Development of the COVID-19 pandemic in the City of Lübeck (above), in Germany (middle) and incidence/100.000 population in the city of Lübeck (below). Sources: City of Lübeck, municipal health department, data for the resident population of Lübeck, status 27/04/2020 at 23:59 hrs; Robert Koch Institute, COVID-19 daily situation reports and dashboard; population data: Destatis; Johns Hopkins University (2020)

A total of 162 patients that were identified as COVID-19 index cases by the local health department were included in the study (Figure 1). Index cases were defined as having acute respiratory symptoms of any severity *or* pneumonia *and* SARS-CoV-2 detection by polymerase chain reaction (PCR)^19^. They were quarantined routinely for at least 14 days from the onset of symptoms or from laboratory testing in the absence of symptoms. After the end of the quarantine period, patients were asked to voluntarily donate blood for antibody testing. Of these 162 index cases, 140 gave their written informed consent to participate and for a total of 110 patients, results from blood samples were available (Fig. 2). Demographic and clinical characteristics of the patients are summarized in Table 1. For children under the age of 18 years parents or other legal guardians provided “informed permission/consent” for study participation. After written consent, serum was analyzed between 3 and 59 days after the end of the quarantine. The patients filled in a questionnaire and subsequently were examined by a GP.

**Figure 2.**
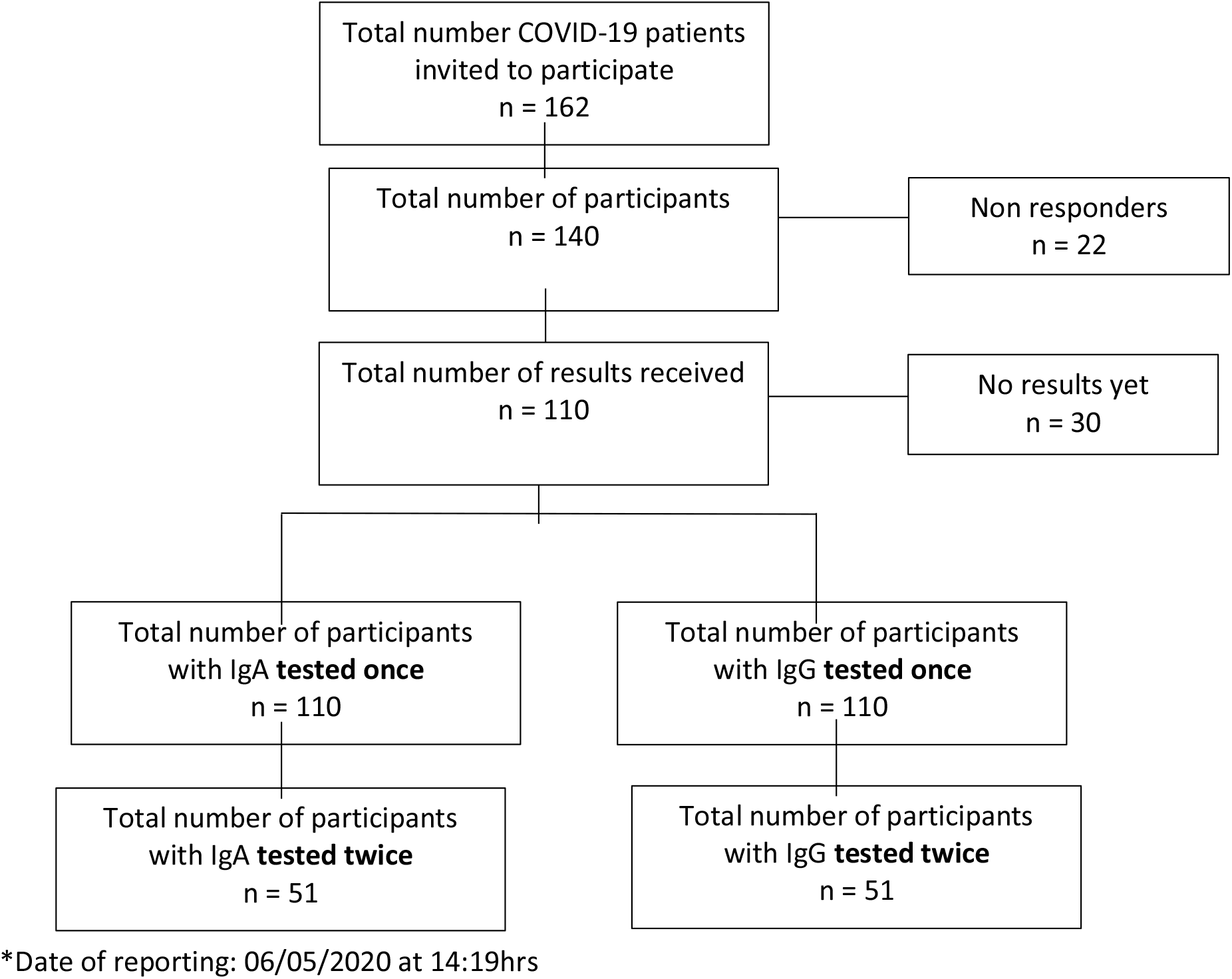
Flowchart of participant enrollment

**Table 1:** Clinical and demographic characteristics of the patients included in the study (n=110)

| Characteristics | (n) (%) |
| --- | --- |
| **Age (median)** | 51 years (16-82) |
| **Gender** |  |
| *Female* | 60 (54.5) |
| *Male* | 50 (45.5) |
| **Disturbance of smell and/or taste** (data available from 96 patients) |  |
| *Yes* | 54 (56.2) |
| *No* | 42 (43.8) |
| **Disease category** (data available from 97 patients) |  |
| 1 *No symptoms* | 10 (10.3) |
| 2. *General weakness, temperature < 38 °C* | 33 (34.0) |
| 3. *General Weakness, dry cough, temperature > 38 °C (influenza like illness)* | 43 (44.3) |
| 4. *As in 3 plus shortness of breath, signs of pneumonia* | 5 (5.2) |
| 5. *As in 4, hospital treatment required* | 6 (6.2) |
| 6. *As in 5, intensive care unit (+- mechanical ventilation)* | 0 |

## Test procedures

### Detection of SARS-CoV-2

Nasopharyngeal swabs were taken from suspected COVID-19 cases by trained professionals either in general practice (GP) or in a “drive-in” swab center run by the municipal health department. Swabs were stored in stabilization media and processed immediately within 4 hours, following DIN EN ISO 17025 und 15189 quality criteria, in the “Laboraerztliche Gemeinschaftspraxis Luebeck", which is located in the immediate vicinity of the “drive-in” swab center. SARS-CoV-2 RNA was detected qualitatively by using an automated one step real-time RT-PCR (RIDA^®^GENE SARS-CoV-2 RUO Test; R-Biopharm AG, Darmstadt, Germany; E-gene amplification) run on a RIDA^®^CYCLER according to the manufacturer’s instruction.

### Detection of anti-SARS-CoV-2 IgG and IgA

Anti-SARS-CoV-2 IgG and IgA were detected by automated enzyme-linked immunosorbent assay - ELISA (product EI 2606-9601 G or A; EUROIMMUN; https://www.euroimmun.com) according to the manufacturer’s instructions. The ELISA plates are coated with recombinant expressed S1 domain glycoprotein of the SARS-CoV-2. According to the latest data sheet (April 2020), the specificity for IgG testing is reported to be 99,1% for IgG and 88,5 % for IgA, respectively. The data sheet reports cross-reactivities with SARS-CoV-1, but not with MERS-CoV, HCoV-229E, HCoV-NL63, HCoVHKU-1 or HCoV-OC43 virus. Thus, possible cross-reactivities are, at most, of marginal importance for this study, since very little, if any, SARS-CoV-1 infection is to be expected.

The optical density (OD) was detected at 450 nm. A ratio of the OD of each sample to the reading of the calibrator, included in the kit, was automatically calculated according to the formula: OD ratio = OD of serum sample/OD of calibrator. According to the manufacturer, a ratio below 0.8 was evaluated as negative, 0.8 - < 1.1 as borderline and >- 1.1 as positive.

### Statistical data

The development of the pandemic in the city of Luebeck was closely monitored by the municipal statistical department based on data from the municipal and national health authorities as well as on own department data. Baseline and demographic characteristics of the patients were summarized by standard descriptive statistics. Since data was not normally distributed, non-parametric tests were chosen. Categorical variables were compared using Fisher’s exact test and continuous variables were compared using the Mann-Whitney U test. Spearman’s correlation was conducted to assess the relation between age and antibody load (IgA and IgG load). Statistical analyses were conducted using SPSS version 26.0. A p-value less than 0.05 was judged statistically significant. The study was designed as a monocentric exploratory study.

## Results

### Local and national development of the COVID-19 pandemic

For the interpretation of our data it is important to first describe the development of the local event. The first two laboratory-confirmed SARS-CoV-2 cases were notified on February 29, 2020, i.e. 41 days after SARS-CoV-2 first reached Germany on January 27. Until March 21, 56 cases were reported to the municipal health department. Fig. 1 shows that from March 21 the number of active cases increased in a linear fashion and reached a plateau-like curve after April 16. From April 1 on, the number of recovered patients always exceeded the active cases. Until May 6th, in total 162 COVID-19 laboratory-confirmed cases and 1 death due to COVID-19 have been reported to the municipal health department of Luebeck.

For Germany, until April 30th, in total 161.539 COVID-19 laboratory-confirmed cases and 6.467 deaths due to COVID-19 were reported. The number for recovered patients exceeded the actively infected cases 13 days later than in the city of Luebeck, i.e. on April 13. After March 16, the local incidence rate always was lower than the national incidence rate (Fig. 1 lower part) and since April 15 was relatively stable at 68.1 – 72.6/100,000, while the incidence in the rest of Germany was steadily increasing (approx. 189/100,000) as of April 27.

### Sample characteristics

As can be seen in Table 1, the median age of the patients was 51 years. Ten patients (9.1 %) were older than 69 years. Sixty (54.5 %) of the patients were female and fifty (45.5 %) were male.

As has been described also by others, a hallmark of COVID-19 disease, like in other viral diseases, is disturbance of smell and/or taste^20^. More than half (56.2 %) of the patients self-reported slight to massive decrease in one or both senses. Although we did not quantify, they reported a duration between one and up to four weeks after recovery from the acute illness. We could not find a significant correlation with age, gender, or disease severity (not shown).

Approx. 10 % of the patients had no symptoms at all. Initially, they were identified as first-degree contact persons through active case finding. After detection of SARS-CoV-2 in nasopharyngeal swabs, they were also classified as index cases. The main symptom in approximately one third of the patients was general weakness with or without headache or bodyache, but no fever (category 2). The age range in this category was between 16 and 80 years. Remarkably, two-thirds (22/33) of the patients in this category were female. 43 out of 97 patients (44.3 %, 22 females, 21 males) had an influenza-like illness with a temperature > 38 °C, in some cases up to 40.2 °C with a reported duration between 15 hours to 72 hours (category 3). They were between 20 and 76 years old. 11 of the patients (6 females, 5 males) had clinical and/or radiological signs of pneumonia. For five of these, hospital treatment was necessary, but no ICU treatment or mechanical ventilation was required. The aged ranged between 22 and 72 years.

### Antibody profiling

162 patients whose quarantine ended between February 29 and April 25, 2020 were eligible for the study. The respective infection time thus was between February 20 and April 11. One day before the end of quarantine, the patients were contacted by phone and asked for their willingness to participate in this study. As of May 6 2020, IgA and IgG antibody results from 110 participants were available. 51 of those were tested twice to monitor the course of antibody development (Fig. 2). Antibodies were analysed between day 10 and day 59 after the end of the quarantine. Figure 3 shows the antibody levels for IgA and IgG in relation to the end of quarantine. The first positive signals were detected on day 10 (i. e. day 24 after SARS-CoV-2 detection) for IgA (Fig. 3a) and for IgG (Fig. 3b). There was a wide inter-individual variation in the antibody levels.

**Figure 3:**
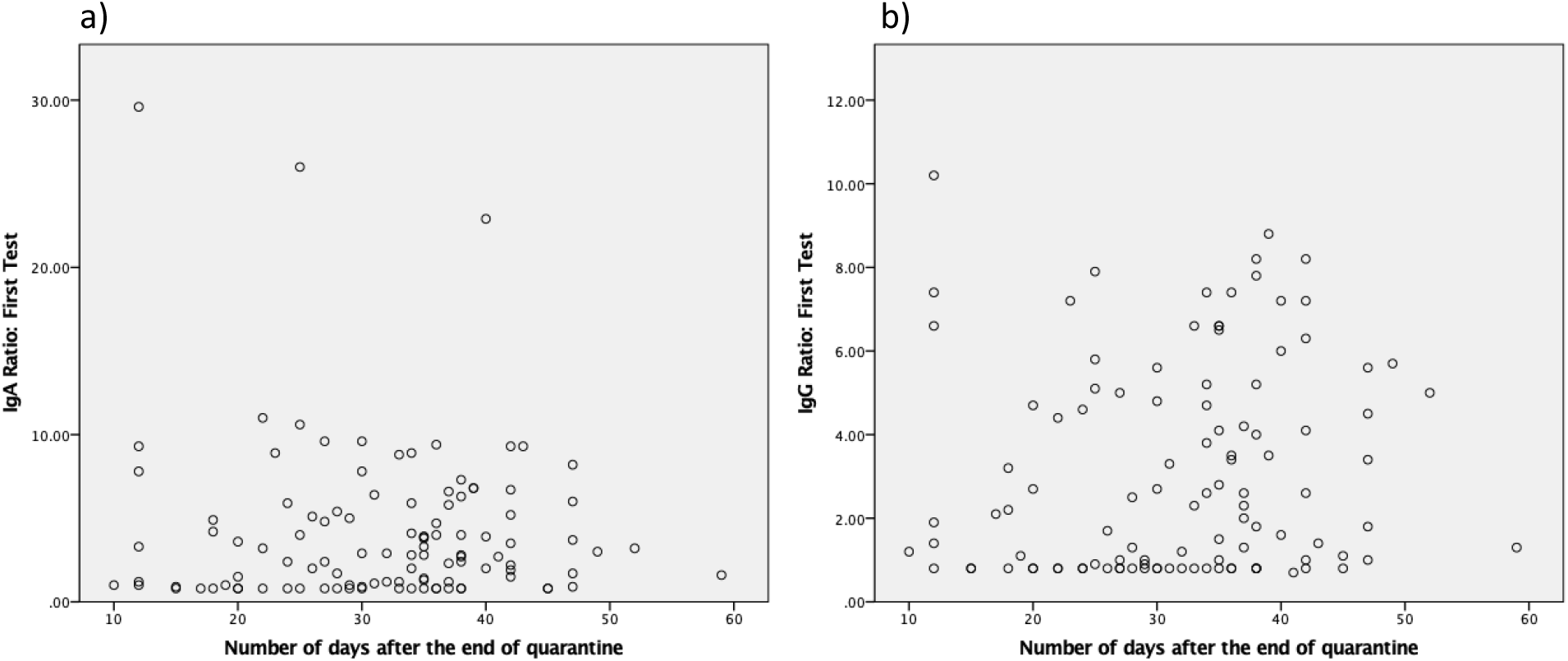
Antibody levels for IgA (a) and IgG (b) in relation to end of quarantine. n= 110

In Table 2, antibody levels from the first and second testing are shown. Given that antibody ratios > 1.1 are defined as positive (according to manufacturer’s data sheet) it is noteworthy that 23.6 % and 29.1 % of the patients were negative for IgA or IgG, respectively (Table 2). It should be remembered here that all patients were tested positive by SARS-CoV-2 PCR. Clinically, most of the seronegative patients were in clinical category 2 or 3, i.e. they had mild to moderate symptoms. On the other hand, eight patients had high IgG antibody levels above 5 in the first testing and 14 patients in a period between 10 and 21 days after the first testing. Again, most of them were in clinical category 2 and 3. No gender difference was seen. When we analysed the 10 asymptomatic patients in our cohort, 6 were antibody negative and two had ratios > 5 (not shown). In 51 patients we analysed a second serum 13, 5 days (range 6 – 21 days) after the first analysis (Fig. 4). It became apparent that for IgA, in most patients the antibody level was lower in the second testing as compared to the first analysis (Fig. 4 upper part). The opposite pattern emerged for IgG (lower part).

**Table 2.** Antibody levels for IgA and IgG

|  | **IgA (1)\***<br>**(n= 110)** | **IgA (2)\*\***<br>**(n= 51)** | **IgG (1)\***<br>**(n= 110)** | **IgG (2)\*\***<br>**(n= 51)** |
| --- | --- | --- | --- | --- |
|  | **N (%)** | **N (%)** | **N (%)** | **N (%)** |
| **< 1.1** | 26 (23.6) | 11 (21.6) | 32 (29.1) | 11 (21.6) |
| **1.1-2** | 20 (18.2) | 16 (31.5) | 22 (20.0) | 16 (31.4) |
| **2.1-5** | 33 (30.0) | 15 (29.4) | 30 (27.3) | 15 (29.4) |
| **5.1-10** | 26 (23.6) | 7 (13.7) | 25 (22.7) | 7 (13.7) |
| **10.1-15** | 2 (1.8) | 1 (2.0) | 1 (0.9) | 1 (2.0) |
| **20.1-25** | 1 (0.6) | 0 | 0 | 0 |
| **>25.1** | 2 (1.2) | 1 (2.0) | 0 | 1 (2.0) |

Note: \*(1) First test \*\* (2) Second test

**Figure 4:**
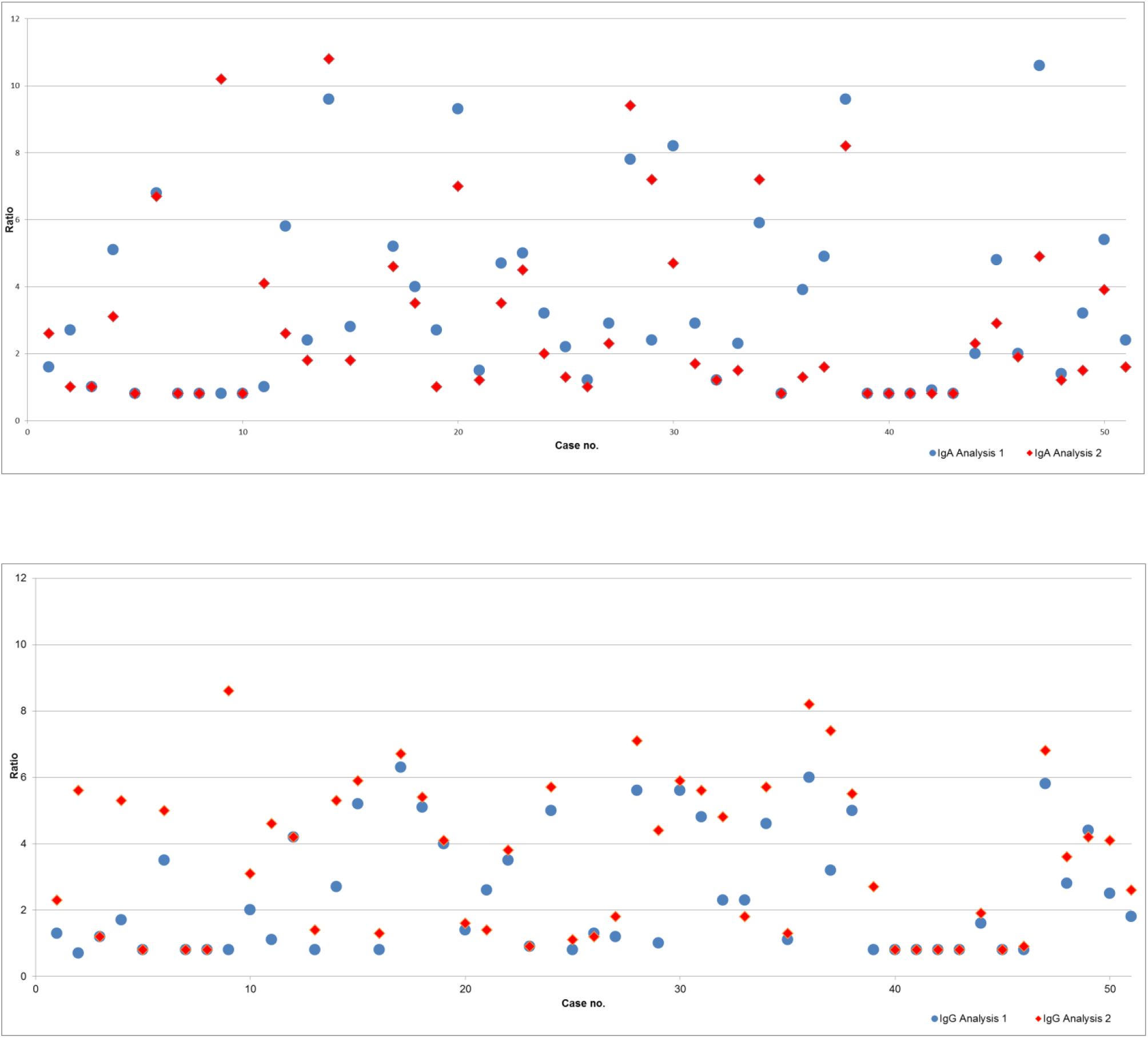
Antibody levels for IgA (upper part) and IgG (lower part) in pairs of serum from individual patients (n=51). The second serum (red) was taken 13.5 days (6 – 21 days) after the first serum (blue).

In summary, in our SARS-CoV-2 PCR-positive cohort, the antibody profile was very heterogeneous. In most cases, antibodies could be detected 10 or more days after the end of the quarantine. Of importance, we could not detect significant antibodies in roughly every fourth patient, despite symptomatic disease. Conversely, 2 out of 10 asymptomatic patients had high antibody levels.

## Discussion

Our analysis of SARS-CoV-2 cases provides insight into the development of immune response in patients with light or moderate course of COVID-19 disease or asymptomatic patients. We analyzed IgA and IgG antibodies recognizing the S1 spike glycoprotein of SARS-CoV-2 in the serum of 110 SARS-CoV-2 PCR-positive patients with mild-to-moderate symptomatic or asymptomatic courses in a region of low-incidence of COVID-19 disease. All patients reside in an area with an incidence of approx. 79 cases/100.000 population in the city of Luebeck. We found a slight dominance of cases in females (54.5 %), which is similar to the recordings for Germany as a whole (52 %)^21^. With regard to initial stratification the course of the disease was mild to moderate. The patients, with a few exceptions, did not require hospital treatment or intensive care unit (ICU) stay. Patients were treated by their family doctors. More than half self-reported sudden disturbances of smell and/or taste with varying durations (days to weeks)^20^. The extent of olfactory dysfunction did not correlate with age or sex or severity of the disease (not shown).

In the analysis of antibody levels, it appeared that, within a period of 50 days after the infection, 84/110 (76 %) and 78/110 patients (71 %) of the patients developed antibodies for IgA and IgG, respectively, above the threshold ratio of 1,1 at the first testing (Table 2). The level of antibodies, however, did not correlate significantly with age or sex or disease severity. Remarkably, 29 % of the patients, with or without symptoms, did not develop IgG antibodies above the cut-off-value (1.1). Again, no correlation with sex, age or disease severity could be observed (not shown).

Ten of the patients did not develop any SARS-CoV-2 specific symptoms. Six out of these did not have detectable antibodies within 50 days. Thus, it is clear that a negative antibody test does not rule out infection. The infectious inoculum possibly was not sufficiently high to induce disease and a subsequent immune reaction. Thus, both, a combination of SARS-CoV-2 PCR and a specific serological test are required to rule out the infection^22^.

Two of the asymptomatic patients had IgG ratios > 5 and the remaining two had levels between 1.1 and 4.9. Although unlikely, it cannot be excluded entirely, that the patients were infected some weeks before the virus was detected and, thus, would have been protected in the observation period of this study. In any case, the data suggest that activation of the humoral immunity might require less virus than activation of symptomatic disease processes. Further studies are needed to define the respective minimal viral loads.

When we investigated the antibody levels in relation to time (Fig. 3), it was obvious that there was a great diversity both for IgA and IgG. For IgA we found some positive values starting at day 10 after the end of quarantine, i.e. 24 days after the infection. It appears, that they decline slowly after day 50. For IgG, the first significant levels (> 1.1) were detected roughly three weeks after the end of quarantine and persisted during the study period. This point of time is later than findings from others suggest^23, 24^ who could detect first antibody titres approx. 2 weeks after the onset of the symptoms. This would correspond to the first day after the end of quarantine in this study. The test systems used were different. It remains to be seen, which antigen (nucleocapsid, spike glycoprotein) is the best to detect relevant antibodies.

We could not find a correlation between antibody levels and age, sex or disease severity (not shown). Based on these findings, it is clear, that antibody diagnosis is a significant pillar to identify COVID-19 - positive patients in addition to SARS-CoV-2 PCR. It also will be indispensable for management of the pandemic. It is likely, that antibodies directed against the S1 glycoprotein are neutralising^17^. For the individual patient, however, it cannot be answered at present, which antibody level will be protective and whether antibody-positive individuals are able to transmit the disease. Further studies are needed to answer this question, which is of utmost importance for example for health care workers. There are few limitations in our study. Cross-reactivity could possibly be a limitation of immunoassays. On the one hand, our test was validated with a sensitivity of 89-100 % and a specificity of 87.5 – 95.5 % for IgA and 83.5 – 97.5 % for IgG and a recent study has demonstrated negligible cross-reactivity from other human coronavirus NL63 to SARS-CoV-2^15, 25^. Our study does not give information on protective antibody functions with regard to resistance to re-infection and reduction of transmissibility of the virus. The results on the neutralisation capacity are not present till now. Based on the development of IgG antibody dynamics, however, it might be reasonable to assume that ratios beyond 2 might confer protection.

## Conclusion

In the present study we could show that approximately three weeks after infection the majority of symptomatic outpatients develops IgA and IgG antibodies in a relevant concentration. In patients with viral RNA detection by PCR, but in the absence of symptoms, significant antibody levels were not detectable in a relevant proportion. This finding raises the question of false-positive PCR results which has to be investigated in further studies. Our data indicate, however, that antibody-positivity is a useful indicator of a previous SARS-CoV-2 infection. Negative antibodies cannot rule out SARS-CoV-2 infection. A number of questions still have to be answered. For the clinic, the determination of the neutralizing capacity of the antibodies in plasma therapy regimes will be of utmost relevance. On a population level, the protective effect for re-infections needs to be determined.

## Data Availability

All relevant data for submission and further analysis are available.

## Ethics

This study was carried out in accordance with the Declaration of Helsinki and the guidelines of the International Conference for Harmonization for Good Clinical Practice. The study was approved by the local Ethics Committee at the University of Luebeck. Informed consent was waived due to the retrospective character of the study. All participants have given their written consent.

## Acknowledgements

The authors would like to thank the patients sharing their data, all medical and laboratory personnel and administrative staff that contributed to obtain the results.

## Abbreviations

COVID-19: coronavirus disease 19
ELISA: enzyme-linked immunosorbent assay
IgG/A: immunoglobulin G/A
rtPCR: real-time polymerase chain reaction
SARS-CoV-2: Severe acute respiratory syndrome coronavirus 2

